# Exploring biomarkers of processing speed and executive function: the role of the anterior thalamic radiations

**DOI:** 10.1101/2022.04.19.22274057

**Authors:** Jennifer Ferris, Brian Greeley, Negin Motamed Yeganeh, Shie Rinat, Joel Ramirez, Sandra Black, Lara Boyd

## Abstract

**Introduction:** Processing speed and executive functioning are often impaired after stroke and in typical aging. However, there are no reliable neurological markers of these cognitive impairments. The trail making test (TMT) is a common index of processing speed and executive function. Here, we tested candidate MRI markers of TMT performance in a cohort of older adults and individuals with chronic stroke.

**Methods:** In 61 older adults and 32 individuals with chronic stroke, we indexed white matter structure with region-specific lesion load (WMH and stroke lesions) and diffusion tensor imaging (DTI) from four regions related to TMT performance: the anterior thalamic radiations (ATR), superior longitudinal fasciculus (SLF), forceps minor, and cholinergic pathways. Regression modelling was used to identify the marker(s) that best predicted TMT performance.

**Results:** DTI metrics of the ATR predicted processing speed in both the older adult (TMT A: β=-3.431, p<0.001) and chronic stroke (TMT A: β=11.282, p<0.001) groups. In the stroke group executive function was best predicted by a combination of ATR and forceps minor DTI metrics in the chronic stroke group (TMT B: _adjusted_R^2^=0.438, p<0.001); no significant predictors of executive function (TMT B) emerged in the older adult group. No imaging metrics related to set shifting (TMT B-A). For all TMT outcome measures with significant imaging predictors, regional DTI metrics predicted TMT performance above and beyond whole-brain stroke and WMH volumes and removing whole-brain lesion volumes improved model fits.

**Conclusions:** In this comprehensive assessment of candidate imaging markers, we demonstrate an association between ATR microstructure and processing speed and executive function performance. Regional DTI metrics provided better predictors of cognitive performance than whole-brain lesion volumes or regional lesion load, emphasizing the importance of lesion location in understanding cognition. We propose ATR DTI metrics as novel candidate imaging biomarker of post-stroke cognitive impairment.

## Introduction

Cognitive impairment affects approximately 40% of individuals who survive a stroke^1^. Specifically, processing speed and executive functions are commonly impaired after stroke^2,3^. Slowed processing speed is associated with poorer functional outcomes^4^, increased dependency^5^, and reduced employment^6^ in individuals with chronic stroke. Despite its high prevalence and clinical importance, we do not understand patterns of damage associated with post-stroke cognitive impairment. To this end, the 2017 consensus meeting of the Stroke Recovery and Rehabilitation Roundtable identified the discovery of brain-based biomarkers of post-stroke cognitive impairment as a research priority^7^.

In addition to stroke infarcts, concurrent aging and vascular neurodegeneration may also impact cognition post-stroke^8^. Declining processing speed and executive function in typical aging are strongly linked to white matter hyperintensities (WMHs)^9^, the most prominent manifestation of cerebral small vessel disease. In typical aging WMHs impact processing speed and executive functions more than any other cognitive domain^10^. Individuals who experience a stroke are more likely to have large WMH volumes^11^, due to the shared cardiometabolic risk factors for large and small vessel cerebrovascular disease^12^. Thus, stroke lesions occur over a background of WMHs, and the common neuropsychological profiles between individuals with stroke and WMHs implies that there may be similar neurobiological pathways that accompany these cognitive changes. Combining information from WMHs and overt stroke lesions could provide new insight into the neurological basis of cognitive impairment post-stroke.

In region-specific analyses of processing speed and executive function four frontal-subcortical white matter tracts have emerged as likely imaging marker candidates: the anterior thalamic radiations (ATR), superior longitudinal fasciculus (SLF), forceps minor of the corpus callosum, and cholinergic pathways of the basal forebrain. The evidence for these regional markers of processing speed and executive function falls into two methodological categories. The first is region-specific lesion load, which measures the degree of overlap between a lesion mask and a white matter tract. WMH lesion load in the ATR^13–15^, SLF^13,14^, and forceps minor^16,17^ relate to processing speed and executive function in older adults. Stroke lesion load in cholinergic pathways relates to processing speed and executive function in individuals with chronic stroke^18^. The second methodological tool is regional microstructural measures obtained from diffusion tensor imaging (DTI). DTI studies have shown some concurrence with regional lesion load studies; processing speed and executive function related to ATR^19,20^ and SLF^19^ microstructure in older adult with WMHs, and SLF microstructure in individuals with subacute stroke^21^. However, in addition to employing different methodological techniques to characterize white matter damage, these previous studies have significant heterogeneity in the types of cognitive assessments used and the regions of interest examined. Thus, it is unknown which specific region (e.g., ATR, SLF, forceps minor, or cholinergic pathways) and which index of structural damage (e.g., lesion load, DTI, or a combination of the two) are the most sensitive predictors of processing speed and executive function.

In this study we indexed processing speed and set shifting, a component of executive function, with the trail making test (TMT), a widely used^22^ and sensitive^23^ neuropsychological assessment. The TMT is composed of two subtests, which are composite cognitive measures. TMT A measures processing speed, including psychomotor speed and visual search^24^. TMT B is more cognitively demanding than TMT A. TMT B relies on higher-order executive functions such as set shifting, in addition to processing speed and visual search components^24^. Additionally TMT B-A can be computed to eliminate the processing speed components in the TMT and specifically index set shifting abilities^25^. The TMT is a useful clinical screening tool; TMT performance predicts conversion from mild cognitive impairment to overt dementia^26^, and TMT B is a predictor of fitness to drive after stroke^27^. We employed the TMT, adapted for a robotic device^28^, to comprehensively test relationships between frontal-subcortical white matter pathways and components of TMT performance.

The goal of the current study was to investigate four candidate brain-based biomarkers of processing speed and executive function (ATR^17,20^, SLF^21,29^, forceps minor^17^, and cholinergic pathways^18^) in a cohort of older adults and individuals in the chronic phase of stroke recovery, across two methodological techniques to assess white matter structure. We indexed structural damage in these pathways using two methods: region-specific lesion load (for both WMH and stroke lesions) and DTI microstructure. We hypothesized that regional white matter markers would explain more variance in TMT performance than whole-brain lesion volumes, but we did not have specific hypotheses about which white matter tracts(s) or structural marker(s) (lesion load vs DTI microstructure) would emerge as the best predictors of TMT performance.

## Methods

### Participants

This study was a secondary analysis of pooled data from the baseline assessments of two research studies conducted by the UBC Brain Behaviour Lab. We included 62 healthy older adults and 34 individuals with chronic stroke who received multimodal neuroimaging between 2016 and 2020. Participants were considered eligible if they were between 40-80 years old, and for the stroke group if they were in the chronic phase of stroke recovery (>6 months post a clinically diagnosed stroke). Participants were ineligible if they: 1) had a history of seizure/epilepsy, head trauma, a major psychiatric diagnosis, neurodegenerative disorders, or substance abuse, or 2) reported any contraindications to MRI. General cognitive performance was assessed with the Montreal Cognitive Assessment (MoCA)^30^. Informed consent was obtained for each participant in accordance with the Declaration of Helsinki. The University research ethics boards approved all aspects of the study protocol.

### Trail making test

All participants completed the TMT on a Kinesiological Instrument for Normal and Altered Reaching Movement (KINARM) end-point device (B-KIN Technologies, Kingston, Ontario). The KINARM TMT is part of the KINARM Standard Tests™ testing battery, and is a robotic adaptation of the pen-and-paper TMT task^30^ (Figure 1). Robotic assessment tools have advantages as objective, reliable and sensitive measures of behaviour^31^. The KINARM endpoint robot requires participants to grip and hold a frictionless manipulandum; this version of the TMT avoids any confound in TMT performance from reduced fine motor control or dexterity^32^

**Figure 1:**
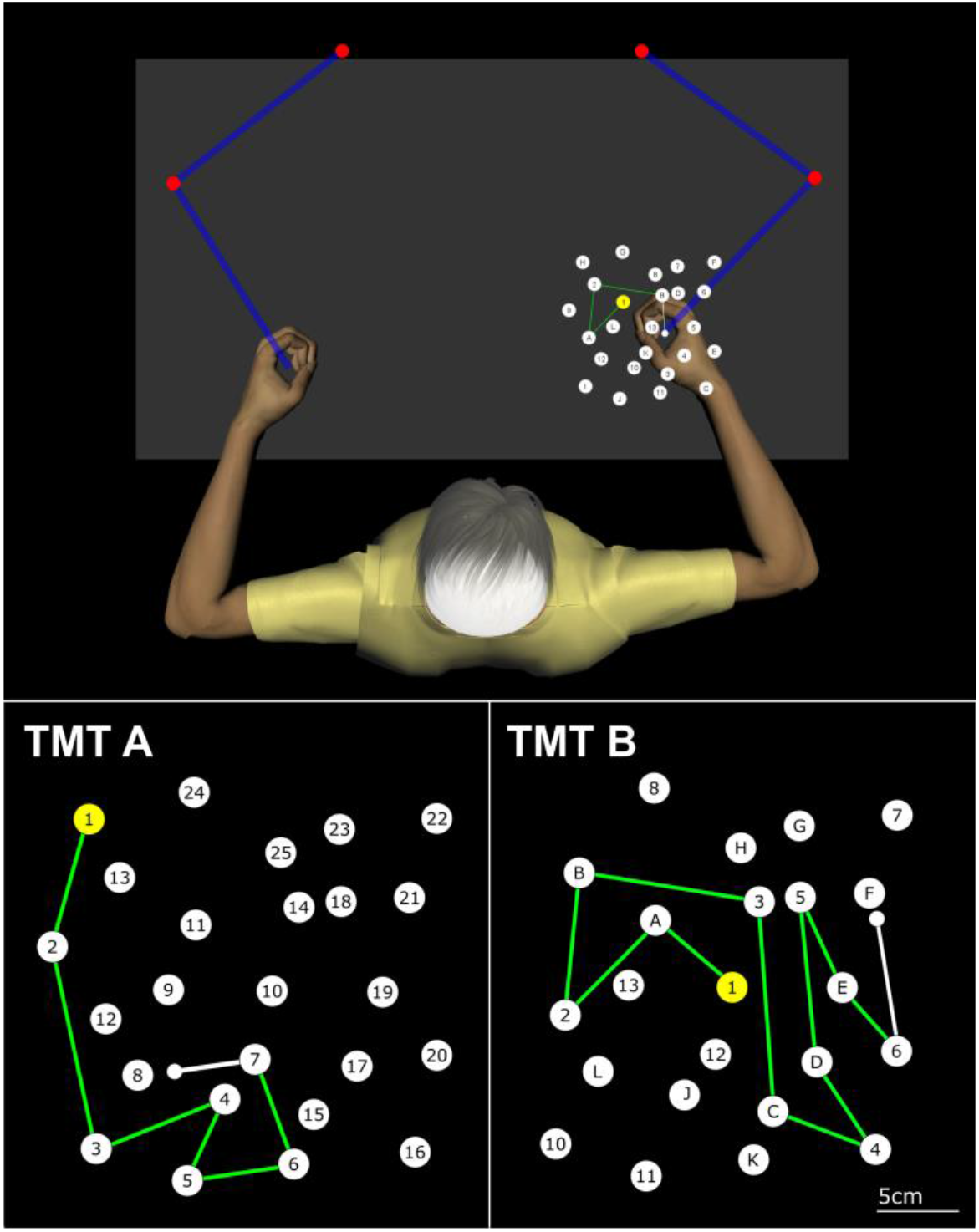
Trail making test experimental setup. Top panel: schematic of the KINARM-adapted trail making task (TMT), showing relative position of the participant and the TMT targets. Participants grabbed an endpoint robotic manipulandum to move their cursor (small white circle) through the TMT targets. Bottom panel: TMT A and B. The start target appeared yellow. Participants moved the cursor between targets, connecting numbers sequentially for TMT A (left) and alternating between numbers and letters in ascending order for TMT B (right). White lines appeared connecting targets mid-movement, which turned green once the participants contacted the next correct target.

Participants were seated, centered in the KINARM workspace. Participants grasped and moved the end-point handles using their dominant (older adult) or non-paretic (stroke) arm during the task, with the hand represented by a small white circle as a cursor. TMT targets were displayed on the dominant/non-paretic side of the KINARM workspace, and participants were instructed to move their hand through the series of targets as quickly and accurately as possible. Regardless of the task, the first target was always numbered 1 and illuminated yellow. As participants moved from one correct target to the next correct target, the line connecting the two targets became fixed and turned green, whereas if participants made an error by moving to an incorrect target, the previously correct target turned red. If this occurred, participants returned to the last correct target (displayed in red) to resume the task. After the instructions and prior to starting the full task, all participants completed a short practice comprised of a five-target version of the task (e.g., 1-2-3-4-5 for TMT A; 1-A-2-B-3 for TMT B).

### Trail Making Task A

The TMT A was comprised of 25 white circular targets numbered 1 to 25 (Figure 1). Targets were randomly distributed in one of 8 possible random patterns. Participants were instructed to connect the targets in ascending numerical sequence as quickly and accurately as possible.

### Trail Making Task B

The TMT B array was comprised of 25 white circular targets with numeric values from 1 to 13 and letter values from A to L (Figure 1). Targets were randomly distributed in one of 8 possible random patterns. Participants were instructed to connect the numbers and letters in ascending alternating alpha-numeric order.

### Outcome measures

We considered three standard TMT outcome measures of interest^33^: total time to complete TMT A, total time to complete TMT B, and TMT B-A time. Total time was defined as the total time taken (seconds) to complete the TMT. The time started once the targets were presented on the screen and ended when the curser contacted the last target. TMT B-A was calculated by subtracting total TMT A time from total TMT B time.

### MRI acquisition

MRI scans were acquired at the University of British Columbia MRI Research Centre on 3.0T Phillips Achieva or Elition scanners (Philips Healthcare, Best, The Netherlands), with parallel imaging and an eight-channel and thirty-two-channel sensitivity encoding head coil, respectively.

We acquired the following structural scans: a 3D magnetization-prepared rapid gradient-echo (MPRAGE) T1 anatomical scan (repetition time (TR)/time to echo (TE)/inversion time (TI) = 3000/3.7/905 ms, flip angle = 9°, voxel size = 1 mm isotropic, field of view (FOV) = 256 × 224 × 180 mm), a fluid attenuated inversion recovery (FLAIR) scan (TR/TE/TI = 9000/90/2500 ms, flip angle = 90°, voxel size = 0.94 × 0.94 mm FOV = 240 × 191 × 144 mm, slice thickness = 3mm), and a combined T2-weighted (T2) and proton density (PD) scan (TR/TE1/TE2 = 2500/9.5/90 ms, flip angle = 90°, voxel size = 0.94 × 0.94 mm, FOV = 240 × 191 × 144mm, slice thickness = 3mm). For DTI data, a high-angular resolution diffusion imaging (HARDI) scan was acquired across 60 non-collinear diffusion gradients (b-value = 700 s/mm^2^, TR/TE = 7094/60ms, voxel size = 2mm isotropic, FOV = 224 × 224 × 154mm, slice thickness = 2.2mm).

### MRI preprocessing

Structural segmentation was performed with the Semi-Automated Brain Region Extraction (SABRE) and Lesion Explorer pipelines^34,35^. Briefly, T1, FLAIR, T2 and PD scans were linearly co-registered and supratentorial cerebral tissue was segmented into cerebrospinal fluid (CSF; sulcal and ventricular), grey matter, normal-appearing white matter (NAWM), and WMHs. Stroke lesions were manually traced over co-registered T1 and FLAIR scans by a single experienced researcher (by J.K.F).

T1 images were skull-stripped using the FMRIB Software Library (FSL) Brain Extraction Tool (BET)^36^. For stroke participants with large cortical lesions, BET often fails to identify the boundaries of the stroke lesion. To improve BET segmentation in the presence of large cortical lesions, stroke lesion masks were set to a voxel intensity roughly corresponding to grey matter and added to T1 scans prior to BET skull strip. The generated binarized BET mask was then used to mask out the original T1 image, to create a skull stripped unaltered T1 image where BET follows the boundaries of the stroke lesion. BET skull strips were visually checked by a single researcher (J.K.F.). T1 scans were non-linearly registered to MNI space using FSL’s FNIRT^37^. To minimize warping in non-linear registration from stroke lesions, stroke lesion masks were flipped across the sagittal midline and a copy of contralesional tissue was created, which was then used to fill in the stroke region on the T1 scan prior to non-linear registration^38^. WMH masks were incorporated in the MNI registration using a cost-function mask, and registration was performed with a warp resolution of 5mm. The quality of MNI registrations was visually confirmed by a single rater (J.K.F.).

Diffusion images were preprocessed with FSL’s diffusion toolbox (FDT)^39^. Briefly, DTI data were corrected for motion and eddy-current distortions, and the unweighted DTI volume was skull-stripped with BET. Fractional anisotropy (FA) and mean diffusivity (MD) maps were generated using DTIFIT. T1 scans were registered to the unweighted DTI volume using FSL’s FLIRT^40^ by a rigid-body linear registration with a correlation ratio cost function. Registration quality was visually checked by a single rater (J.K.F.) and manually adjusted where necessary with tkregister from Freesurfer v.6.0.

### Regions of interest

ATR, SLF, and forceps minor tracts were taken as regions of interest (ROIs) from the JHU white matter atlas^41^ and binarized with a probability threshold of 0.1^13,42^. The left and right hemispheres of ATR and SLF were combined to create a single bilateral ROI for each region. To calculate regional lesion load: WMH and stroke masks were moved to MNI space, and we calculated the overlap between the lesion mask and the ROI. We then computed a weighted lesion load for each ROI, according to previously published methods^43^. For lesion load calculations, ROIs were sliced in the plane that corresponded to capturing the cross-sectional area of each ROI (coronal plane for ATR and SLF; sagittal plane for forceps minor). To calculate regional DTI metrics: ROIs were moved to T1 space and eroded to subject-specific white matter anatomy by removing voxels containing grey matter or cerebrospinal fluid. Next, ROIs were moved to DTI space and mean FA and MD were extracted. Based on the Cholinergic Pathways HyperIntensities Scale (CHIPS)^44^, cholinergic pathways were automatically parcellated in T1 space from the SABRE atlas, and WMH and stroke lesion load was calculated in the cholinergic pathways, according to previously published methods^45^. DTI measures were not extracted from cholinergic pathways because this area contains unmyelinated white matter fibers^46^ and overlaps with the trajectory of the SLF^18^.

### Statistics

Statistical analyses were performed with R (programming environment v4.0.4); the alpha threshold for significance was set at *p* < 0.05. Whole-brain lesion volumes and tract-specific lesion load measures were positively skewed and were log-transformed prior to statistical analysis. Relationships between imaging markers and TMT performance were tested separately for each group (older adults, chronic stroke), and outcome measure (TMT A, TMT B, and TMT B-A). First, we performed exploratory Spearman’s correlations between cognitive outcome measures and imaging metrics (WMH lesion load, stroke lesion load, FA, and MD) for each ROI (ATR, SLF, forceps minor, and cholinergic pathways). Next, we entered any variables that significantly correlated with the TMT outcome measure into linear regression models, to test which regional markers, or combination of markers, best predicted TMT performance. Regression models were adjusted for age and time post stroke (for chronic stroke group models). Scanner was added as an additional control variable in the older adult models to account for potential effects. All predictor variables were mean centered and standardized. To identify which imaging metrics best predicted cognitive performance, we performed model testing of linear regression models by removing the predictor with the smallest beta-value, and comparing model fit with adjusted R^2^ and Akaike’s information criterion (AIC). We tested for collinearity in predictors by calculating the variance inflation factor (VIF). Separate models were constructed if VIF > 3.0 for any predictors^47^. Final models in this regression analysis were selected if the imaging metric was a significant predictor in the final model, the overall regression model was significant, and the model gave the highest R^2^ and lowest AIC in the tested model sets. We estimated effect sizes of predictors in the model with partial *η*^2^.

Once final models were established for TMT performance, we tested whether region-specific imaging metrics or whole-brain lesion volumes explained more variance in TMT performance. We fit a model entering both whole-brain lesion volumes and imaging metrics as predictors. Next, we compared performance between models fit with only whole-brain lesion volumes or regional imaging metrics as predictors.

Finally, we compared model performance between imaging data separated by cerebral hemisphere (left versus right hemisphere for older adult group, ipsilesional versus contralesional for chronic stroke group) to test whether data from one hemisphere explained greater variance in TMT performance. For this analysis we excluded participants with bilateral stroke lesions. We further interrogated the role of hemisphere in TMT performance with a supplementary analysis of individuals with chronic stroke, where we tested relationships between hemispheric imaging data and TMT performance between individuals with left hemisphere versus right hemisphere lesions.

## Results

We included 62 otherwise healthy older adults and 34 individuals with chronic stroke in this analysis. All participants were successfully processed through MRI processing steps. One older adult was missing DTI data, but their lesion load and TMT data was included. One older adult and two individuals with chronic stroke were considered outliers in TMT times (≥3 standard deviations (SDs) above the mean); data from these individuals were excluded from final analyses. Our final sample included 61 older adults (age range: 46 - 80 years old, 23 males, 56 right-hand dominant) and 32 individuals with chronic stroke (age range: 45 - 80 years old, 22 males, 7 bilateral stroke infarcts, 13 left hemisphere stroke infarcts, 12 right hemisphere stroke infarcts). In the older adult group, 39 individuals were scanned on the Phillips Achieva MRI, and 22 were scanned on the Phillips Elition MRI.

Table 1 presents participant demographics and mean TMT data for each group. Lesion overlap images for WMHs and stroke lesions are presented in Supplementary Figure 1. Relative to older adults, the individuals with chronic stroke had lower MoCA scores, larger whole-brain WMH volumes, and a lower proportion of females in the sample. Individuals with chronic stroke also had poorer performance on the TMT indicated by longer total TMT A and TMT B time, and greater differences in time to complete (TMT B-A). Table 2 presents mean imaging metrics between groups. Relative to older adults, individuals with chronic stroke had significantly lower FA and higher MD in ATR, SLF, and forceps minor (Table 2: all p < 0.001), and significantly greater WMH lesion load in ATR, SLF, forceps minor and cholinergic pathways (Table 2: all p < 0.010). We tested for potential sex differences in imaging metrics, presented in Supplementary Table 1. There were no differences in any imaging metrics between sexes (all p > 0.05).

**Table 1:**
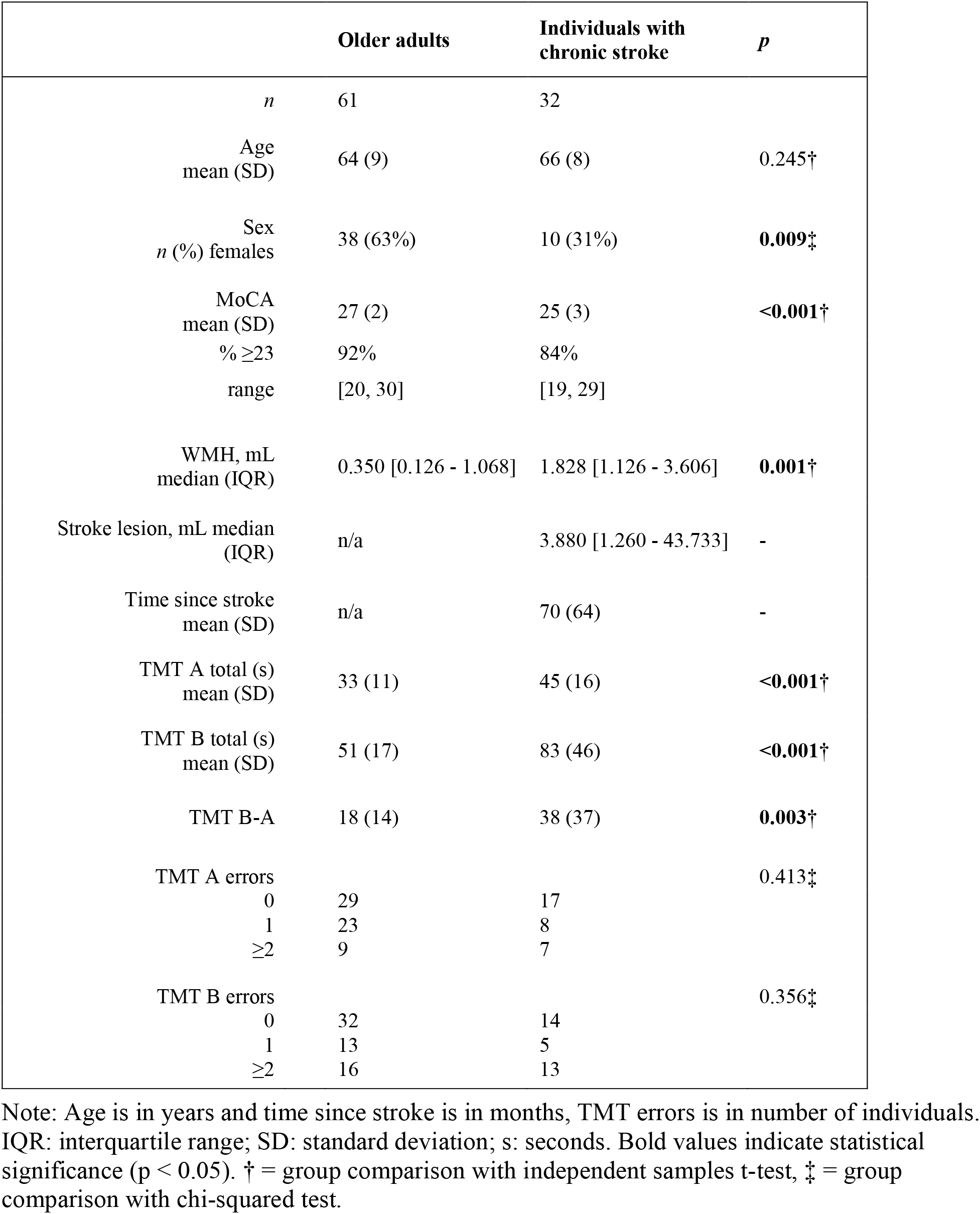
Participant demographics.

|  | Older adults | Individuals with chronic stroke | *p* |
| --- | --- | --- | --- |
| *n* | 61 | 32 |  |
| Age<br>mean (SD) | 64 (9) | 66 (8) | 0.245<sup>†</sup> |
| Sex<br>*n* (%) females | 38 (63%) | 10 (31%) | **0.009**<sup>‡</sup> |
| MoCA<br>mean (SD) | 27 (2) | 25 (3) | **<0.001**<sup>†</sup> |
| % $\geq 23$ | 92% | 84% | |
| range | [20, 30] | [19, 29] |  |
| WMH, mL<br>median (IQR) | 0.350 [0.126 - 1.068] | 1.828 [1.126 - 3.606] | **0.001**<sup>†</sup> |
| Stroke lesion, mL median<br>(IQR) | n/a | 3.880 [1.260 - 43.733] | - |
| Time since stroke<br>mean (SD) | n/a | 70 (64) | - |
| TMT A total (s)<br>mean (SD) | 33 (11) | 45 (16) | **<0.001**<sup>†</sup> |
| TMT B total (s)<br>mean (SD) | 51 (17) | 83 (46) | **<0.001**<sup>†</sup> |
| TMT B-A | 18 (14) | 38 (37) | **0.003**<sup>†</sup> |
| TMT A errors |  |  | 0.413<sup>‡</sup> |
| 0 | 29 | 17 |  |
| 1 | 23 | 8 |  |
| $\geq 2$ | 9 | 7 | |
| TMT B errors |  |  | 0.356<sup>‡</sup> |
| 0 | 32 | 14 |  |
| 1 | 13 | 5 |  |
| $\geq 2$ | 16 | 13 | |

Note: Age is in years and time since stroke is in months, TMT errors is in number of individuals. IQR: interquartile range; SD: standard deviation; s: seconds. Bold values indicate statistical significance ( $p < 0.05$ ). <sup>†</sup> = group comparison with independent samples t-test, <sup>‡</sup> = group comparison with chi-squared test.

**Table 2:**
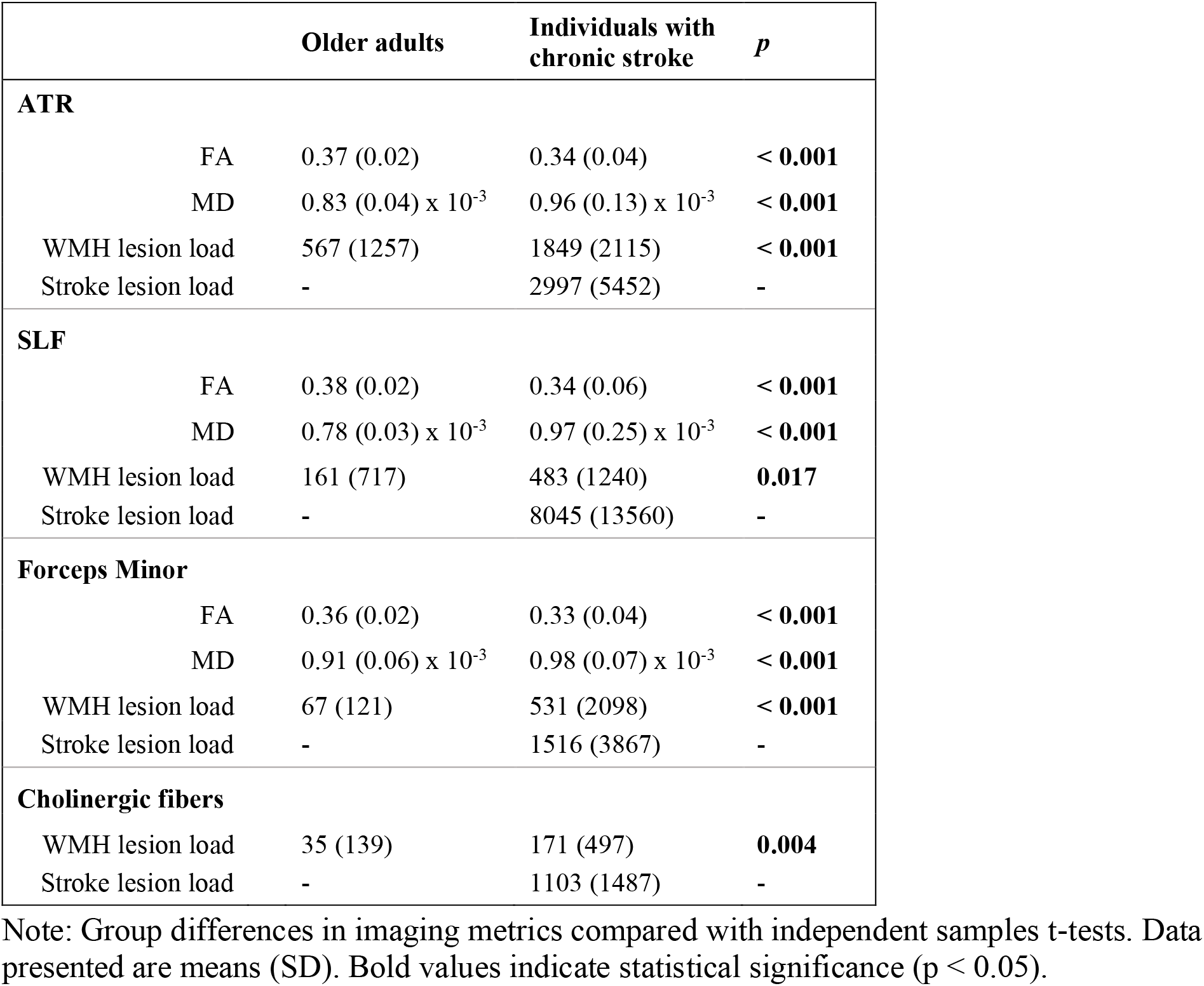
Group differences in imaging metrics.

### TMT models in older adults

Bivariate spearman’s correlations revealed that total TMT A time significantly correlated with the following variables: ATR FA (r_s_ = −0.410, p = 0.001), forceps minor FA (r_s_ = −0.335, p = 0.009), SLF MD (r_s_ = 0.335, p = 0.009), SLF FA (r_s_ = −0.289, p = 0.025), and ATR WMH lesion load (r_s_ = 0.279, p = 0.029). These variables were therefore included in linear regression model testing.

Full regression model testing is presented for older adults in Table 3. The best linear regression model fit was achieved by ATR FA as a predictor, which showed a negative relationship with TMT A time after accounting for age and MRI scanner (Table 3). All other imaging variables failed to account for a significant amount of variance in TMT A time, and model performance was improved after these variables were removed.

**Table 3:**
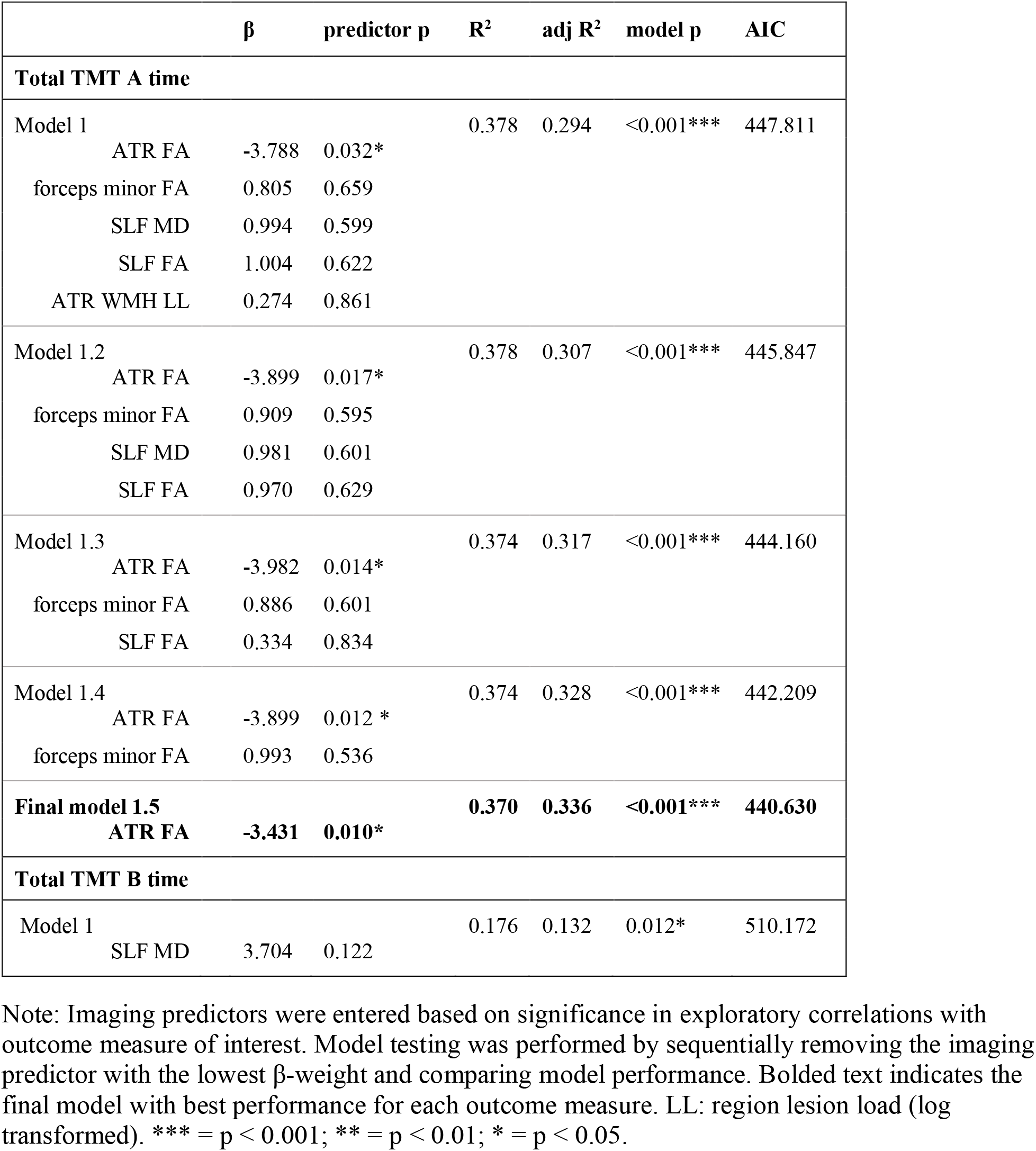
Imaging predictors of TMT performance in older adults.

| | $\beta$ | predictor p | R<sup>2</sup> | adj R<sup>2</sup> | model p | AIC |
| --- | --- | --- | --- | --- | --- | --- |
| **Total TMT A time** |  |  |  |  |  |  |
| Model 1 |  |  | 0.378 | 0.294 | <0.001*** | 447.811 |
| ATR FA | -3.788 | 0.032* |  |  |  |  |
| forceps minor FA | 0.805 | 0.659 |  |  |  |  |
| SLF MD | 0.994 | 0.599 |  |  |  |  |
| SLF FA | 1.004 | 0.622 |  |  |  |  |
| ATR WMH LL | 0.274 | 0.861 |  |  |  |  |
| Model 1.2 |  |  | 0.378 | 0.307 | <0.001*** | 445.847 |
| ATR FA | -3.899 | 0.017* |  |  |  |  |
| forceps minor FA | 0.909 | 0.595 |  |  |  |  |
| SLF MD | 0.981 | 0.601 |  |  |  |  |
| SLF FA | 0.970 | 0.629 |  |  |  |  |
| Model 1.3 |  |  | 0.374 | 0.317 | <0.001*** | 444.160 |
| ATR FA | -3.982 | 0.014* |  |  |  |  |
| forceps minor FA | 0.886 | 0.601 |  |  |  |  |
| SLF FA | 0.334 | 0.834 |  |  |  |  |
| Model 1.4 |  |  | 0.374 | 0.328 | <0.001*** | 442.209 |
| ATR FA | -3.899 | 0.012 * |  |  |  |  |
| forceps minor FA | 0.993 | 0.536 |  |  |  |  |
| **Final model 1.5** |  |  | **0.370** | **0.336** | **<0.001\*\*\*** | **440.630** |
| **ATR FA** | **-3.431** | **0.010\*** |  |  |  |  |
| **Total TMT B time** |  |  |  |  |  |  |
| Model 1 |  |  | 0.176 | 0.132 | 0.012* | 510.172 |
| SLF MD | 3.704 | 0.122 |  |  |  |  |

Note: Imaging predictors were entered based on significance in exploratory correlations with outcome measure of interest. Model testing was performed by sequentially removing the imaging predictor with the lowest $\beta$ -weight and comparing model performance. Bolded text indicates the final model with best performance for each outcome measure. LL: region lesion load (log transformed). \*\*\* = $p < 0.001$ ; \*\* = $p < 0.01$ ; \* = $p < 0.05$ .

Bivariate spearman’s correlations revealed that total TMT B time was significantly correlated with SLF MD (r_s_ = 0.387, p = 0.002), but SLF MD did not remain a significant predictor of TMT B time in the linear regression model after accounting for age and MRI scanner (Table 3). TMT B-A time did not significantly correlate with any imaging metrics (all p > 0.05) and therefore did not progress to regression model testing.

### TMT models in individuals with chronic stroke

Bivariate spearman’s correlations (r_s_) revealed that total TMT A time significantly correlated with the following variables: ATR FA (r_s_ = −0.460, p = 0.008), ATR MD (r_s_ = 0.431, p = 0.014), and forceps minor stroke lesion load (r_s_ =0.355, p = 0.046). These variables were therefore included in linear regression model testing.

Full regression model testing is presented for individuals with chronic stroke in Table 4. Model testing was performed across two separate models due to high collinearity between ATR FA and MD data (VIF > 4.0). The best linear regression model fit was achieved by ATR MD as a predictor, which showed a negative relationship with total TMT A time, after accounting for age and time since stroke (Table 4). ATR FA was also significantly related to total TMT A time, but the ATR FA model explained less variance in TMT A time than the ATR MD model (adjusted (adj) R^2^ ATR FA model: 0.273, vs. adj R^2^ ATR MD model: 0.390).

**Table 4:** Imaging predictors of TMT performance in individuals with chronic stroke.

| | | $\beta$ | predictor p | $R^2$ | adj $R^2$ | model p | AIC |
| --- | --- | --- | --- | --- | --- | --- | --- |
| **Total TMT A time** |  |  |  |  |  |  |  |
| Model 1 |  |  |  | 0.413 | 0.326 | 0.005** | 261.797 |
|  | ATR FA | -8.424 | 0.004** |  |  |  |  |
|  | forceps minor stroke LL | 5.064 | 0.085 |  |  |  |  |
| Model 1.1 |  |  |  | 0.343 | 0.273 | 0.007** | 263.372 |
|  | ATR FA | -10.053 | <0.001*** |  |  |  |  |
| Model 2 |  |  |  | 0.452 | 0.371 | 0.002** | 259.567 |
|  | ATR MD | 12.305 | 0.002** |  |  |  |  |
|  | forceps minor stroke LL | -1.514 | 0.689 |  |  |  |  |
| **Final Model 2.2** |  |  |  | **0.449** | **0.390** | **< 0.001\*\*\*** | **257.760** |
|  | **ATR MD** | **11.282** | **<0.001\*\*\*** |  |  |  |  |
| **Total TMT B time** |  |  |  |  |  |  |  |
| **Final Model 1** |  |  |  | **0.511** | **0.438** | **<0.001\*\*\*** | **324.441** |
|  | **ATR FA** | **-40.485** | **<0.001\*\*\*** |  |  |  |  |
|  | **forceps minor FA** | **14.740** | **0.065** |  |  |  |  |
| Model 1.1 |  |  |  | 0.444 | 0.384 | < 0.001*** | 326.542 |
|  | ATR FA | -31.406 | <0.001*** |  |  |  |  |
| Model 2 |  |  |  | 0.372 | 0.279 | 0.011* | 332.411 |
|  | ATR MD | 29.180 | 0.010* |  |  |  |  |
|  | forceps minor MD | -1.814 | 0.858 |  |  |  |  |
| Model 2.2 |  |  |  | 0.371 | 0.304 | 0.004** | 330.449 |
|  | ATR MD | 27.844 | <0.001*** |  |  |  |  |
| **TMT B-A** |  |  |  |  |  |  |  |
|  | forceps minor MD | 11.525 | 0.088 | 0.166 | 0.076 | 0.161 | 325.721 |

Note: Imaging predictors were entered based on significance in exploratory correlations with outcome measure. Model testing was performed by sequentially removing the imaging predictor with the lowest $\beta$ -weight and comparing model performance. Bolded text indicates the final model with best performance for each TMT outcome measure. LL: region lesion load (log transformed). \*\*\* = $p < 0.001$ ; \*\* = $p < 0.01$ ; \* = $p < 0.05$ .

Bivariate spearman’s correlations revealed that total TMT B time was significantly correlated with the following variables: forceps minor MD (r_s_ = 0.550, p = 0.001), ATR FA (r_s_ = − 0.521, p = 0.002), ATR MD (r_s_ = 0.477, p = 0.006), forceps minor FA (r_s_ = −0.376, p = 0.034). These variables were included in linear regression model testing, across two separate models due to high collinearity between FA and MD data (VIF > 4.0). The best linear regression model fit was achieved with ATR FA and forceps minor FA as imaging predictors (Table 4). ATR MD was also a significant predictor of total TMT B time, but the ATR MD model explained less variance in TMT B time than the ATR & forceps minor FA model (adj R^2^ ATR & forceps minor FA model: 0.438 vs adj R^2^ ATR MD model: 0.304).

TMT B-A time was significantly correlated with forceps minor MD (r_s_ = 0.500, p = 0.004), but forceps minor MD was not a significant predictor of TMT B-A in the linear regression model after accounting for age and time post-stroke (Table 4).

### Region-specific markers versus whole-brain lesion volumes

We tested whether region-specific imaging markers explained more variance in TMT performance compared to whole-brain lesion volumes. We performed model comparison only for TMT measures with significant predictors in the previous linear regression model testing (total TMT A time for older adult group, total TMT A & B time for chronic stroke group). Full model comparisons are presented in Table 5. For every tested TMT outcome measure, regional DTI metrics explained a significant amount of variance in TMT performance over and above whole-brain lesion volumes. Further, the best fitting model in each case was the model with regional DTI metrics only, which provided a better fit to the data then models with whole-brain lesion volumes alone or models with combined whole-brain lesion volumes and regional DTI metrics (see Table 5). In other words, regional DTI metrics explained TMT variance above and beyond whole-brain lesion volumes, and model performance was improved when whole-brain lesion volumes were dropped.

**Table 5:**
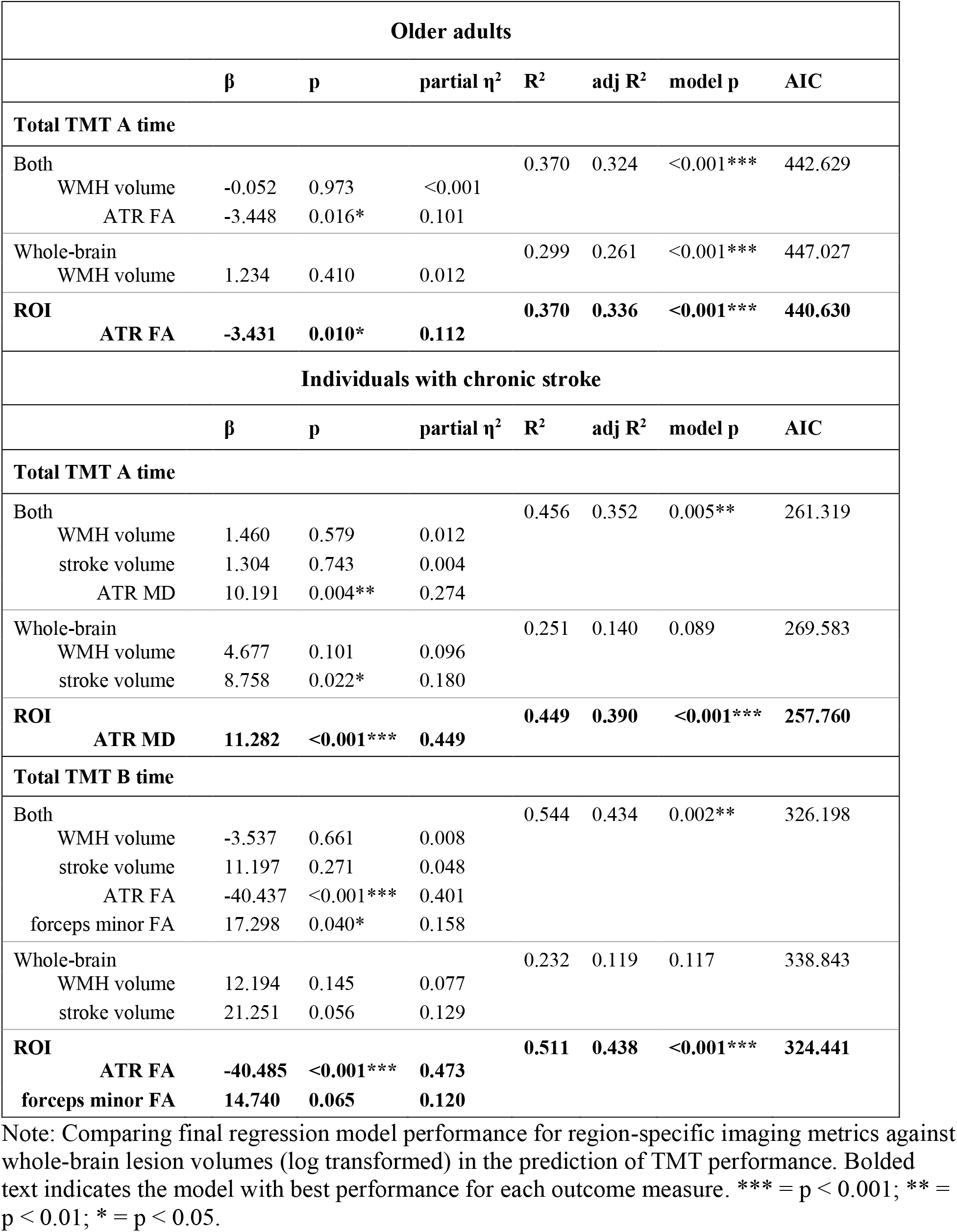
Regional imaging metrics versus whole-brain lesion volumes to predict TMT performance.

| Older adults |  |  |  |  |  |  |  |
| --- | --- | --- | --- | --- | --- | --- | --- |
| | $\beta$ | p | partial $\eta^2$ | $R^2$ | adj $R^2$ | model p | AIC |
| **Total TMT A time** |  |  |  |  |  |  |  |
| Both |  |  |  | 0.370 | 0.324 | <0.001*** | 442.629 |
| WMH volume | -0.052 | 0.973 | <0.001 |  |  |  |  |
| ATR FA | -3.448 | 0.016* | 0.101 |  |  |  |  |
| Whole-brain |  |  |  | 0.299 | 0.261 | <0.001*** | 447.027 |
| WMH volume | 1.234 | 0.410 | 0.012 |  |  |  |  |
| **ROI** |  |  |  | **0.370** | **0.336** | **<0.001\*\*\*** | **440.630** |
| **ATR FA** | **-3.431** | **0.010\*** | **0.112** |  |  |  |  |
| **Individuals with chronic stroke** |  |  |  |  |  |  |  |
| | $\beta$ | p | partial $\eta^2$ | $R^2$ | adj $R^2$ | model p | AIC |
| **Total TMT A time** |  |  |  |  |  |  |  |
| Both |  |  |  | 0.456 | 0.352 | 0.005** | 261.319 |
| WMH volume | 1.460 | 0.579 | 0.012 |  |  |  |  |
| stroke volume | 1.304 | 0.743 | 0.004 |  |  |  |  |
| ATR MD | 10.191 | 0.004** | 0.274 |  |  |  |  |
| Whole-brain |  |  |  | 0.251 | 0.140 | 0.089 | 269.583 |
| WMH volume | 4.677 | 0.101 | 0.096 |  |  |  |  |
| stroke volume | 8.758 | 0.022* | 0.180 |  |  |  |  |
| **ROI** |  |  |  | **0.449** | **0.390** | **<0.001\*\*\*** | **257.760** |
| **ATR MD** | **11.282** | **<0.001\*\*\*** | **0.449** |  |  |  |  |
| **Total TMT B time** |  |  |  |  |  |  |  |
| Both |  |  |  | 0.544 | 0.434 | 0.002** | 326.198 |
| WMH volume | -3.537 | 0.661 | 0.008 |  |  |  |  |
| stroke volume | 11.197 | 0.271 | 0.048 |  |  |  |  |
| ATR FA | -40.437 | <0.001*** | 0.401 |  |  |  |  |
| forceps minor FA | 17.298 | 0.040* | 0.158 |  |  |  |  |
| Whole-brain |  |  |  | 0.232 | 0.119 | 0.117 | 338.843 |
| WMH volume | 12.194 | 0.145 | 0.077 |  |  |  |  |
| stroke volume | 21.251 | 0.056 | 0.129 |  |  |  |  |
| **ROI** |  |  |  | **0.511** | **0.438** | **<0.001\*\*\*** | **324.441** |
| **ATR FA** | **-40.485** | **<0.001\*\*\*** | **0.473** |  |  |  |  |
| **forceps minor FA** | **14.740** | **0.065** | **0.120** |  |  |  |  |

Note: Comparing final regression model performance for region-specific imaging metrics against whole-brain lesion volumes (log transformed) in the prediction of TMT performance. Bolded text indicates the model with best performance for each outcome measure. \*\*\* = $p < 0.001$ ; \*\* = $p < 0.01$ ; \* = $p < 0.05$ .

### Hemisphere-specific effects

We tested whether observed TMT relationships with imaging markers were driven by hemisphere effects. For this analysis, we excluded seven participants with bilateral stroke infarcts, leaving a sample size of 25 in the chronic stroke group. We performed model comparison unilateral ATR tracts against TMT measures with significant predictors in the previous linear regression model testing (total TMT A time for older adult group, total TMT A and B time for chronic stroke group). Full model comparisons are presented in Table 6. For the older adult group, both right and left hemisphere ATR FA were significant predictors of TMT A performance, however right hemisphere ATR FA explained more variance in the TMT A data, by a small margin (Table 6; adj R^2^ right hemisphere model: 0.336 vs. adj R^2^ left hemisphere model: 0.324). For the chronic stroke group, both TMT A and TMT B time were predicted by ATR data from the ipsilesional, but not contralesional, hemisphere (Table 6).

**Table 6:** Hemispheric effects of imaging predictors of TMT performance.

| Older adults |  |  |  |  |  |  |  |
| --- | --- | --- | --- | --- | --- | --- | --- |
| | $\beta$ | p | partial $\eta^2$ | R<sup>2</sup> | adj R<sup>2</sup> | model p | AIC |
| **TMT A** |  |  |  |  |  |  |  |
| Left hemisphere<br>ATR FA | -3.112 | 0.017* | 0.097 | 0.359 | 0.324 | <0.001*** | 441.641 |
| **Right hemisphere**<br>**ATR FA** | **-3.528** | **0.010\*\*** | **0.113** | **0.370** | **0.336** | **<0.001\*\*\*** | **440.566** |
| **Individuals with chronic stroke** |  |  |  |  |  |  |  |
| | $\beta$ | p | partial $\eta^2$ | R<sup>2</sup> | adj R<sup>2</sup> | model p | AIC |
| **TMT A** |  |  |  |  |  |  |  |
| Contralesional hemisphere<br>ATR MD | 7.046 | 0.063 | 0.155 | 0.164 | 0.045 | 0.278 | 217.961 |
| **Ipsilesional hemisphere**<br>**ATR MD** | **15.968** | **<0.001\*\*\*** | **0.521** | **0.526** | **0.459** | **0.001\*\*** | **203.770** |
| **TMT B** |  |  |  |  |  |  |  |
| Contralesional hemisphere<br>ATR FA | -26.620 | 0.027* | 0.222 | 0.314 | 0.177 | 0.095 | 268.309 |
| forceps minor FA | 3.895 | 0.733 | 0.006 |  |  |  |  |
| **Ipsilesional hemisphere**<br>**ATR FA** | **-41.665** | **0.002\*\*** | **0.385** | **0.458** | **0.350** | **0.012\*** | **262.419** |
| **forceps minor FA** | **7.998** | **0.430** | **0.031** |  |  |  |  |

Note: Comparing final regression model performance between hemispheres for unilateral tracts in prediction of TMT performance. Bolded text indicates the model with best performance for each predictor variable. \*\*\* = $p < 0.001$ ; \*\* = $p < 0.01$ ; \* = $p < 0.05$ .

To further interrogate the role of lesioned hemisphere in the observed DTI-TMT relationships, we conducted a supplementary analysis comparing TMT performance and model fits between individuals with right hemisphere strokes (n = 12) and left hemisphere strokes (n = 13). TMT performance was not different between individuals with right and left hemisphere strokes (Supplementary Table 2; all p > 0.05). Within each group, TMT performance was best predicted by the ipsilesional hemisphere, regardless of whether the ipsilesional hemisphere was a right or left hemispheres lesion (Supplementary Table 3).

## Discussion

In this study we tested regional white matter predictors of processing speed and set shifting indexed by performance on the TMT. We evaluated multiple frontal-subcortical white matter tracts (ATR, SLF, forceps minor, and cholinergic pathways) using two methods to quantify structural damage in these tracts (regional lesion load and DTI microstructure). Our study has three main findings. First, DTI metrics of the ATR emerged as the best predictor of TMT performance for both older adults and individuals with chronic stroke. Second, ATR DTI metrics explained significantly more variance in TMT performance than stroke or WMH volumes from the whole-brain. Third, ipsilesional, but not contralesional, ATR data related to TMT performance in individuals with chronic stroke, whereas both dominant and non-dominant ATR related to TMT performance in older adults.

The development of a cognitive biomarker could facilitate advances in cognitive rehabilitation; a promising, but understudied, area of rehabilitation^48^. This study provides encouraging preliminary evidence for ATR microstructure as a candidate biomarker of processing speed and, to a lesser degree, executive function. For context, in the chronic stroke group the effect size of ipsilesional ATR MD with TMT A performance (partial η^2^ = 0.526; Table 6) is comparable to effect sizes seen for ipsilesional corticospinal tract FA and Fugl-Meyer scores (range 0.51-0.62^49^), a widely accepted neuroimaging marker of upper extremity motor outcomes post-stroke^7^.

### Trail making test and cognition

The TMT is a commonly used cognitive assessment for both healthy and clinical populations and measures multiple components of cognitive performance. For TMT A, we found that higher ATR FA was associated with faster times to complete the TMT A for both older adults and individuals with chronic stroke, although in the chronic stroke group ATR MD explained more variance in TMT A times than ATR FA. We are the first study to show a relationship between ATR microstructure and processing speed in individuals with chronic stroke.

TMT B performance in individuals with chronic stroke was best predicted by a combination of FA from the ATR and forceps minor. Individuals with agenesis of the corpus callosum show deficits in TMT B task performance^50^, which may indicate a contribution of interhemispheric communication to the cognitive processes underlying TMT B performance. In our older adult group TMT B performance was not significantly predicted, which may be explained by the characteristics of our sample. Our older adult sample had very low WMH volumes (mean 1.25 mL, roughly corresponding to a Fazekas score ≤1^51^) and relatively intact cognitive functioning (average MoCA score was 27, 92% of the sample scored greater than 23 on the MoCA^52^). Previous studies reporting relationships between ATR structure and TMT B performance in older adults included participants with significantly greater WMH load than the current older adult sample^17,20^. Our data are in line with the hypothesis that critical structures for cognitive function become evident only with greater cumulative damage to white matter tracts^17^ as we observed relationships between ATR microstructure and TMT B performance in the chronic stroke group only.

No white matter tracts emerged as significant predictors of TMT B-A. TMT B-A was the most precise cognitive measure in our battery, and as such it may also require more specific neuroanatomical localization. fMRI studies suggest set-shifting is associated with dorsolateral prefrontal cortex (DLPFC) activity^53,54^. The ATR region encompasses all fibers of the anterior limb of the internal capsule; while this includes projections between the thalamus and DLPFC^55^, it also captures projections to other frontal cortex regions including from the medial forebrain bundle^56^. It may be that relationships between thalamocortical structure and set-shifting ability will emerge with individualized tractography between the thalamus and DLPFC.

Our findings suggest a relatively greater contribution of ATR microstructure to processing speed rather than set shifting, as ATR microstructure related to TMT A performance across our sample and predicted TMT B performance in the chronic stroke group, whereas TMT B-A, a specific measure of set shifting, did not have any significant predictors. Additionally, individuals with chronic stroke took longer to complete TMT-B but did not make more errors relative to the older adult group. This indicates that it is slowed speed, rather than increased set-shifting errors, that contributed to TMT-B performance differences between groups. This argument is supported by Stuss *et al*. who reported that patients with focal frontal lobe lesions and executive dysfunction made more errors on TMT-B, but did not show differences in raw TMT-B completion time^25,57^. A major theory of aging suggests that slowed processing speed is a central process that underlies poor performance in multiple other cognitive domains^58^. Our data extend this theory, suggesting that frontal thalamocortical circuitry may be an early mediator of slowed processing speed. Considering the importance of processing speed to multiple components of cognitive abilities^58^, our findings underscore the potential utility of ATR microstructure for use as a brain-based biomarker of post-stroke cognitive impairment.

### Anterior thalamic radiation

The ATR connects the thalamus and the frontal cortex via the anterior limb of the internal capsule^41^. The ATR is sensitive to age-related degeneration; in a study of 3,500 adults Cox *et al*. found that the ATR had the largest age-related changes in FA and MD among 14 white matter tracts^59^. Similarly, among thalamic grey matter nuclei, regions with frontal cortex connectivity show the most atrophy with aging^60^. These age-related findings might reflect the vulnerability of the ATR to damage from WMHs, because the anterior limb of the internal capsule travels through the periventricular region of the lateral ventricles, the most common site of WMH formation^61^. The vulnerability of the ATR to WMH formation would explain both the prevalence of processing speed and executive function deficits seen in individuals with WMHs^10^, and the structure-function relationships observed in the current study. ATR therefore is a promising candidate marker of cognitive performance, given its widespread connectivity with the frontal lobes^41^, its vulnerability to age and vascular related degeneration^59^, and its relationships with cognitive performance in this and previous^13–15,19,20^ reports.

### DTI-based biomarkers

In this study we employed two methodological indexes of white matter structure to explain variability in TMT performance: DTI and regional lesion load. DTI markers, but not regional lesion load markers, related to TMT performance. ATR DTI microstructure was consistently the best predictor across all tested regions and structural metrics, and it predicted TMT performance above and beyond whole-brain WMH and stroke volumes. Conversely, region-specific lesion load metrics did not survive regression model testing, and all final models included only DTI metrics.

DTI likely emerged as a more sensitive structural index because DTI can capture more variability in white matter structure than regional lesion volumetrics. In addition to underlying tract anatomy, DTI microstructure is sensitive to both stroke lesions^62^ and WMHs^63^. Furthermore, DTI can detect subtle changes to white matter that extend beyond the boundaries of a segmented lesion, such as in the so-called “WMH-penumbra”^63–65^ and in stroke-related Wallerian degeneration along the length of tracts^66^. In contrast, lesion volumetrics are based on a binary measure of categorizing a voxel as lesioned or non-lesioned, and thus may miss more subtle effects of lesions on tract structure. In support of this idea, we found that DTI data from the ipsilesional ATR drove relationships with TMT performance, implying that stroke-related damage is involved in the relationship between ATR and TMT performance. However, stroke lesion load in the ATR did not relate to TMT performance, indicating DTI is capturing variability in ATR microstructure above and beyond the stroke lesion that contributes to TMT decline, potentially from co-occurring WMHs. DTI has promise as a clinical tool because it provides a sensitive index of multiple forms of brain damage, and therefore may be useful across a variety of clinical populations. Future research should continue to interrogate the interactions between overt stroke lesions and WMH on behavioural outcomes.

## Limitations

Our study was cross-sectional in design; given this we cannot test whether ATR microstructure predicts individual trajectories of cognitive decline. While the cross-sectional design served to explore candidate markers of processing speed and executive functions, future longitudinal studies are needed to evaluate the predictive potential of ATR microstructure for cognitive performance. Second, we employed an atlas-based approach to white matter pathway delineation, which cannot capture individual differences in white matter anatomy. We balanced this limitation by eroding atlas regions to individual white matter anatomy. An atlas-based approach has several strengths, notably it is more easily scalable for clinical translation. However, future studies could use individualistic tractography approaches, especially to test contributions from specific cortical targets such as DLPFC thalamocortical pathways. Finally, it is difficult to disentangle the contributions of processing speed and executive function in cognitive tasks^67^, and future studies should more fully investigate the cognitive functions supported by ATR microstructure with a full neuropsychological testing battery.

## Conclusion

Amongst a group of neuroimaging candidates, ATR microstructure emerged as a robust predictor of TMT A performance in older adults and individuals with chronic stroke, and TMT B performance in individuals with chronic stroke. TMT A and TMT B are complex tests that rely on multiple cognitive abilities; however, our data suggest a relatively greater contribution of ATR structure to processing speed, compared to set shifting abilities. ATR microstructure related to TMT performance above and beyond whole-brain WMH and stroke volumes, indicating the importance of regional metrics when considering behavioural relationships. ATR microstructure is a promising candidate for the development of novel markers to predict cognitive decline and response to intervention in cerebrovascular disease.

## Data Availability

All data produced in the present study are available upon reasonable request to the corresponding author.

## Acknowledgements

We gratefully acknowledge the assistance of imaging analysts in the LC Campbell Cognitive Neurology Research Unit, including Sabrina Adamo, Miracle Ozzude, Dr. Fuqiang Gao, and Christopher Scott. We thank Dr. Teresa Liu Ambrose for helpful conversations related to this paper. Funding was provided by the Canadian Institutes of Health Research (MOP-130269, PTJ-148535 and PTJ-153330; PI Boyd).

