## Supplementary material for "Exploring biomarkers of processing speed and executive function: the role of the anterior thalamic radiations": Data Supplement

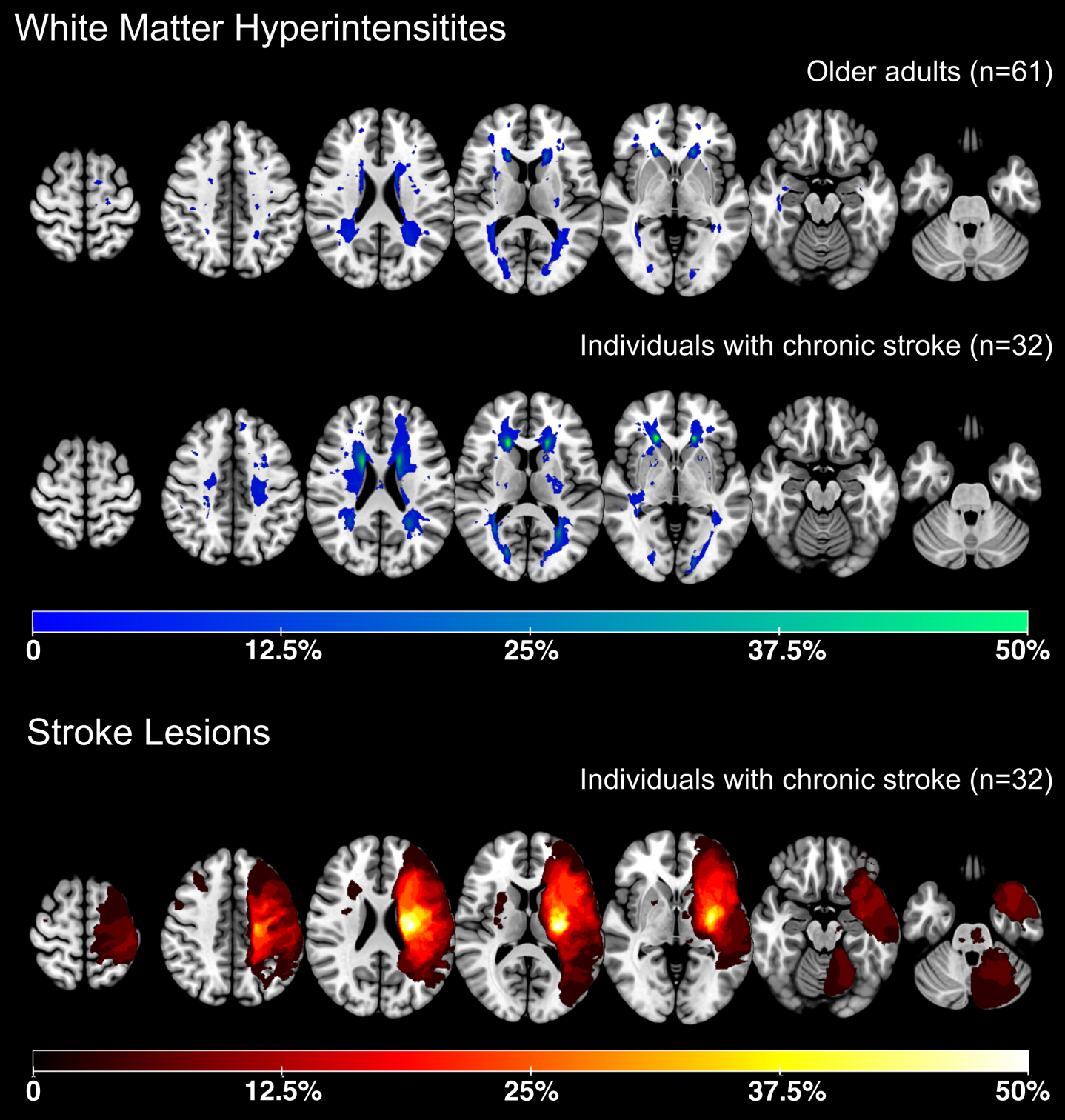

Supplementary Figure 1: Lesion overlap images for white matter hyperintensities (WMH; in blue) and chronic stroke lesions (in red). WMH maps are split by group (older adults and individuals with chronic stroke). Stroke lesions were flipped along the L/R axis so that all symptomatic strokes (contralateral to the impaired upper extremity) were visualized in the left hemisphere. Color bar represents the percentage of the sample with a lesion in each voxel.

|  |  | **Older adults** | **Individuals with chronic stroke** | ***p*** |
| --- | --- | --- | --- | --- |
| **ATR** |  |  |  |  |
| FA |  | 0.37 (0.02) | 0.34 (0.04) | **< 0.001** |
| MD |  | 0.83 (0.04) x 10^-3^ | 0.96 (0.13) x 10^-3^ | **< 0.001** |
| WMH lesion load |  | 567 (1257) | 1849 (2115) | **< 0.001** |
| Stroke lesion load |  | - | 2997 (5452) | **-** |
| **SLF** |  |  |  |  |
| FA |  | 0.38 (0.02) | 0.34 (0.06) | **< 0.001** |
| MD |  | 0.78 (0.03) x 10^-3^ | 0.97 (0.25) x 10^-3^ | **< 0.001** |
| WMH lesion load |  | 161 (717) | 483 (1240) | **0.017** |
| Stroke lesion load |  | - | 8045 (13560) | **-** |
| **Forceps Minor** | | | | |
| FA |  | 0.36 (0.02) | 0.33 (0.04) | **< 0.001** |
| MD |  | 0.91 (0.06) x 10^-3^ | 0.98 (0.07) x 10^-3^ | **< 0.001** |
| WMH lesion load |  | 67 (121) | 531 (2098) | **< 0.001** |
| Stroke lesion load |  | - | 1516 (3867) | - |
| **Cholinergic fibers** | | | | |
| WMH lesion load |  | 35 (139) | 171 (497) | **0.004** |
| Stroke lesion load |  | - | 1103 (1487) | - |

Supplementary Table 1: Group differences in imaging metrics, compared with independent samples t-tests. ATR: Anterior thalamic radiation; FA: Fractional anisotropy; MD: mean diffusivity; SLF: superior longitudinal fasciculus, WMH: white matter hyperintensity. Bold values indicate statistical significance (p < 0.05)

|  |  | **Females  (n = 38)** | **Males  (n = 23)** | ***p*** |
| --- | --- | --- | --- | --- |
| **WMH volume, mL** |  | 1.41 (3.33) | 0.97 (1.77) | 0.556 |
| **ATR** |  |  |  |  |
| FA |  | 0.37 (0.02) | 0.38 (0.02) | 0.140 |
| MD |  | 0.83 (0.05) x 10^-3^ | 0.83 (0.04) x 10^-3^ | 0.949 |
| WMH lesion load |  | 4.82 (2.18) | 3.96 (2.65) | 0.171 |
| **SLF** |  |  |  |  |
| FA |  | 0.38 (0.02) | 0.38 (0.03) | 0.677 |
| MD |  | 0.78 (0.04) x 10^-3^ | 0.79 (0.03) x 10^-3^ | 0.336 |
| WMH lesion load |  | 1.98 (2.50) | 0.89 (1.89) | 0.075 |
| **Forceps Minor** |  |  |  |  |
| FA |  | 0.36 (0.03) | 0.37 (0.03) | 0.609 |
| MD |  | 0.91 (0.07) x 10^-3^ | 0.91 (0.05) x 10^-3^ | 0.994 |
| WMH lesion load |  | 2.97 (1.84) | 2.36 (2.11) | 0.245 |
| **Cholinergic fibers** |  |  |  |  |
| WMH lesion load |  | 1.18 (1.87) | 0.60 (1.59) | 0.234 |

Supplementary Table 2: Sex differences in imaging metrics for older adults, compared with independent samples t-tests.

|  |  | **Females  (n = 10)** | **Males  (n = 22)** | ***p*** |
| --- | --- | --- | --- | --- |
| WMH volume, mL |  | 4.14 (5.88) | 3.91 (4.89) | 0.906 |
| Stroke volume, mL |  | 24.80 (55.34) | 34.18 (49.10) | 0.633 |
| **ATR** |  |  |  |  |
| FA |  | 0.33 (0.04) | 0.34 (0.03) | 0.208 |
| MD |  | 0.96 (0.14) x 10^-3^ | 0.95 (0.13) x 10^-3^ | 0.843 |
| WMH lesion load |  | 7.01 (1.27) | 6.65 (1.44) | 0.498 |
| Stroke lesion load |  | 4.89 (3.11) | 5.12 (3.55) | 0.863 |
| **SLF** |  |  |  |  |
| FA |  | 0.34 (0.07) | 0.34 (0.06) | 0.637 |
| MD |  | 0.93 (0.25) x 10^-3^ | 0.98 (0.26) x 10^-3^ | 0.527 |
| WMH lesion load |  | 3.61 (2.60) | 2.53 (2.69) | 0.296 |
| Stroke lesion load |  | 5.58 (3.42) | 5.43 (4.39) | 0.927 |
| **Forceps Minor** |  |  |  |  |
| FA |  | 0.33 (0.05) | 0.33 (0.03) | 0.851 |
| MD |  | 0.96 (0.07) x 10^-3^ | 0.98 (0.07) x 10^-3^ | 0.479 |
| WMH lesion load |  | 4.51 (1.29) | 4.42 (2.00) | 0.893 |
| Stroke lesion load |  | 1.30 (3.04) | 1.90 (3.42) | 0.636 |
| **Cholinergic fibers** |  |  |  |  |
| WMH lesion load |  | 2.44 (2.63) | 2.14 (2.16) | 0.740 |
| Stroke lesion load |  | 5.99 (2.29) | 4.13 (3.54) | 0.138 |

Supplementary Table 3: Sex differences in imaging metrics for individuals with chronic stroke, compared with independent samples t-tests.

|  |  | **Individuals with left hemisphere strokes** | **Individuals with right hemisphere strokes** | ***p*** |
| --- | --- | --- | --- | --- |
| Total TMT A time  mean (SD) |  | 47 (20) | 43 (15) | 0.569 |
| Total TMT B time  mean (SD) |  | 99 (59) | 73 (37) | 0.192 |
| TMT B-A  mean (SD) |  | 53 (42) | 30 (35) | 0.162 |

Supplementary Table 4: Differences in trail making test (TMT) performance between individuals with left (n = 13) and right (n = 12) hemisphere strokes, compared with independent samples t-tests.

| **Individuals with left hemisphere strokes  (n = 13)** | | | | | | | | |
| --- | --- | --- | --- | --- | --- | --- | --- | --- |
|  |  | **β** | **p** | **partial η^2^** | **R^2^** | **adj R^2^** | **model p** | **AIC** |
| **TMT A** |  |  |  |  |  |  |  |  |
| **Left hemisphere** |  |  |  |  | **0.556** | **0.407** | **0.054** | **112.906** |
| **ATR MD** |  | **18.696** | **0.009**** | **0.546** |  |  |  |  |
| Right hemisphere |  |  |  |  | 0.415 | 0.220 | 0.167 | 116.479 |
| ATR MD |  | 13.448 | 0.036* | 0.402 |  |  |  |  |
| **TMT B** |  |  |  |  |  |  |  |  |
| **Left hemisphere** |  |  |  |  | **0.632** | **0.449** | **0.065** | **140.662** |
| **ATR FA** |  | **-58.597** | **0.008**** | **0.602** |  |  |  |  |
| **Forceps minor FA** |  | **12.071** | **0.443** | **0.075** |  |  |  |  |
| Right hemisphere |  |  |  |  | 0.377 | 0.066 | 0.377 | 147.508 |
| ATR FA |  | -42.530 | 0.084 | 0.327 |  |  |  |  |
| Forceps minor FA |  | 1.278 | 0.947 | 0.001 |  |  |  |  |
| **Individuals with right hemisphere strokes (n = 12)** | | | | | | | | |
|  |  | **β** | **p** | **partial η^2^** | **R^2^** | **adj R^2^** | **model p** | **AIC** |
| **TMT A** |  |  |  |  |  |  |  |  |
| Left hemisphere |  |  |  |  | 0.097 | -0.241 | 0.834 | 106.315 |
| ATR MD |  | 4.929 | 0.596 | 0.037 |  |  |  |  |
| **Right hemisphere** |  |  |  |  | **0.510** | **0.326** | **0.111** | **98.988** |
| **ATR MD** |  | **13.806** | **0.027*** | **0.477** |  |  |  |  |
| **TMT B** |  |  |  |  |  |  |  |  |
| Left hemisphere |  |  |  |  | 0.524 | 0.252 | 0.211 | 122.687 |
| ATR FA |  | -21.936 | 0.495 | 0.069 |  |  |  |  |
| Forceps minor FA |  | 8.184 | 0.631 | 0.035 |  |  |  |  |
| **Right hemisphere** |  |  |  |  | **0.616** | **0.397** | **0.111** | **120.103** |
| **ATR FA** |  | **-22.409** | **0.171** | **0.249** |  |  |  |  |
| **Forceps minor FA** |  | **10.224** | **0.427** | **0.092** |  |  |  |  |

Supplementary Table 5: Comparing final model performance between groups of individuals with left hemisphere strokes, compared to right hemisphere strokes. Bolded text indicates the model with best performance for each predictor variable. Regardless of which hemisphere was affected by the stroke, the ipsilesional hemisphere explained more variance in TMT performance than the contralesional hemisphere (i.e., the left hemisphere for individuals with left hemisphere lesions, and the right hemisphere for individuals with right hemisphere lesions). ** = p < 0.01; * = p < 0.05.
